# Implementing remote monitoring for COVID-19 patients in primary care

**DOI:** 10.1101/2024.02.27.24303073

**Authors:** Svea Holtz, Susanne M. Köhler, Peter Jan Chabiera, Nurlan Dauletbaev, Kim Deutsch, Zoe Oftring, Dennis Lawin, Lukas Niekrenz, Teresa Euler, Rainer Gloeckl, Rembert Koczulla, Gernot Rohde, Michael Dreher, Claus F. Vogelmeier, Sebastian Kuhn, Beate Sigrid Müller

## Abstract

**Background:** In Germany, most patients with coronavirus disease 2019 (COVID-19) are treated in an outpatient setting. To improve assessments of the health status of COVID-19 outpatients, various remote monitoring models have been developed. However, little information exists on experiences acquired with remote monitoring in an outpatient setting, particularly from a patient perspective. The aim of our ‘COVID-19@home’ study was therefore to implement and evaluate an app-based remote monitoring concept for acute and post-acute COVID-19-patients in primary care. In this paper, we focus on the patients’ evaluation of our remote monitoring approach.

**Methods:** Patients with acute COVID-19 measured heart rate, blood pressure, oxygen saturation, and body temperature daily for 28 days. Patients with post-acute COVID-19 determined the same parameters for 12 weeks, supplemented by lung parameters and daily step count. The data were documented using the ‘SaniQ’ smartphone app. COVID-19 symptoms were assessed daily using an app-based questionnaire. Patients’ GPs could access the data on the ‘SaniQ Praxis’ telemedicine platform. We used an app-based questionnaire consisting of 11 questions presented with a 4-point Likert scale to evaluate patient satisfaction. Data were analyzed descriptively.

**Results:** Of the 51 patients aged 19-77 years that participated in the study, 42 completed the questionnaire. All patients rated home monitoring as ‘very good’ or ‘rather good’ and were able to integrate the measuring processes into their daily routines. Overall, 93% would recommend the app and the measuring devices to their family and friends. About 60% felt that their COVID-19 treatment had benefited from home monitoring. Only few patients were unsettled by the app and use of the measuring devices. During the course of the study, the implementation process was optimized.

**Conclusions:** The use of remote monitoring in COVID-19 patients is feasible and was evaluated positively by most study patients. However, it is difficult to imagine how general practices could cope with monitoring patients with acute diseases without any further organizational support. Future research should address cost-effectiveness and changes in such clinical outcomes as hospitalization and mortality.

## 1. Introduction

In Germany, the majority of patients with a severe acute respiratory syndrome coronavirus 2 (SARS-CoV-2) infection are treated in an outpatient setting. This was true for 6/7 patients in the first phase of the pandemic and thereafter for 32/33 patients, as a result of milder disease courses in vaccinated patients [1]. While most patients present with mild or moderate symptoms [2], some develop severe clinical outcomes and require hospitalization. Despite receiving such treatment as invasive mechanical ventilation, or kidney replacement therapy, some of these patients may die [3]. The early identification of patients showing preliminary signs of deterioration is therefore crucial, since early interventions, including hospitalization, are associated with better outcomes and prognoses [4]. Furthermore, the severity of symptoms may not adequately reflect organ damage, especially as coronavirus disease 2019 (COVID-19) is sometimes associated with ‘silent hypoxia’, an abrupt and initially asymptomatic drop in oxygen saturation [5, 6]. Standardized monitoring of blood oxygen saturation is expected to enable the timely recognition of patients requiring treatment escalation [6–8].

To better assess the health status of outpatients with COVID-19, health care providers in several countries reacted to the global outspread of SARS-CoV-2 by applying remote monitoring solutions [9, 10]. Some providers, for example, used (video) phone-calls to gather information, while others developed mobile applications (apps) to collect data on vital signs [9]. However, both general practitioners and patients have little experience of using remote monitoring for acute diseases [11].

We therefore initiated the ‘Covid-19@home’ study, with the aim of implementing and evaluating a remote monitoring concept for COVID-19-patients, i.e. patients with an acute disease, in primary care. We used the ‘SaniQ’ app and the corresponding ‘SaniQ Praxis’ telemonitoring platform [12]. In this paper, we focus on participating patients’ adherence and their evaluation of our approach to remote monitoring, as well as the optimization of the implementation process.

## 2. Methods

### Study design and setting

We conducted a prospective observational study (COVID-19@home) in Frankfurt, Germany. The study was carried out from January to December 2021, whereby the period from

January 2021 to July 2021 was a pilot phase to optimize the implementation process. The data presented in this study were collected between January and November 2021. For this paper, we followed STROBE reporting guidelines for cohort studies, where applicable [13].

### Study registration and ethical approval

COVID-19@home was part of the egePan Unimed project, and one of the thirteen projects that make up ‘Netzwerk Universitätsmedizin’ [14]. It was registered in the German Clinical Trials Register (DRKS00024604, https://www.drks.de/drks_web/setLocale_EN.do, 26.04.2021). The study was approved by the Ethics Committee of Goethe University Frankfurt (No. 20-1023, 18.01.2021), and written informed consent was obtained from all participants.

### GPs

Eight GPs participated in the study. Seven of them belonged to a research network of general practices in Hesse, Germany (ForN) and were recruited after participating in a virtual network meeting. One GP agreed to participate after receiving an invitation to participate by mail. All GPs provided written informed consent prior to study participation. Qurasoft GmbH, Koblenz, Germany, provided each GP with his or her own password-protected user account for version 1.x of the SaniQ Praxis telemonitoring platform free of charge. Before recruiting patients, each GP attended a 30-minute one-on-one online training session on how to use the platform. The session was conducted by the software company Qurasoft GmbH using Microsoft Teams software. GPs did not receive any compensation for their participation.

### Patients

Participating GPs asked patients that had had a PCR test for COVID-19 at their practice if they wished to participate. They were included in the study if their PCR test result was positive (acute COVID-19 patients). Patients that were being treated by their GPs for persistent COVID-19 symptoms (post-acute COVID-19 patients) were also asked if they wanted to participate. Patients had to be aged ≥ 18 years and have access to a smartphone that they, or someone in the same household, was able to use. GPs decided which patients to invite. They handed out a flyer that contained information on the study, and the phone number and email address of the study team. Patients could then contact the study team via telephone or email if they were interested in participating. The study team at the Institute of General Practice at Goethe University Frankfurt answered all questions by telephone.

Patients received detailed written study information and were required to give their written informed consent. They were also informed that neither the study team nor the treating GP provided 24/7-supervision, and that they were required to monitor their vital signs themselves. In case of medical queries, patients were advised to call their GPs or the emergency services. They were further required to confirm that they had understood these conditions on the consent form. Study participation was voluntary and could be terminated at any time. The patients received no monetary compensation but were permitted to keep the measuring devices.

### Measuring devices and patient app

The patients received the required measuring equipment within 48 hours of inclusion in the study, so they could quickly begin monitoring their vital signs. Acute COVID-19 patients received a pulse oximeter, a blood pressure monitor, and a non-contact thermometer, while post-acute patients additionally received a peak flow and FEV1 meter, and an activity sensor (see table 1). Patients were also provided with two codes to activate and authorize the smartphone app ‘SaniQ’ (module ‘Infekt’). All equipment was provided free of charge. After installing the app and entering the activation code, the patients used the devices to measure their health parameters. Data were entered into the app manually, or transmitted from the devices via Bluetooth®. The app was used to document the measured data and patients could view the data in the app (raw data and graphs), enabling them to monitor the course of the disease themselves. When a health parameter fell outside the normal range, the app sent an in-app notification or email to the patient (alert) stating that, for example, ‘The measured value for your heart rate is outside the normal range. Please repeat the measurement. In case of uncertainty, contact your GP practice, or the emergency services.’ During business hours, the study team could be reached on the telephone to answer questions that were unrelated to health. Additionally, the app and platform provider (Qurasoft GmbH) provided a hotline for technical problems.

**Table 1.**
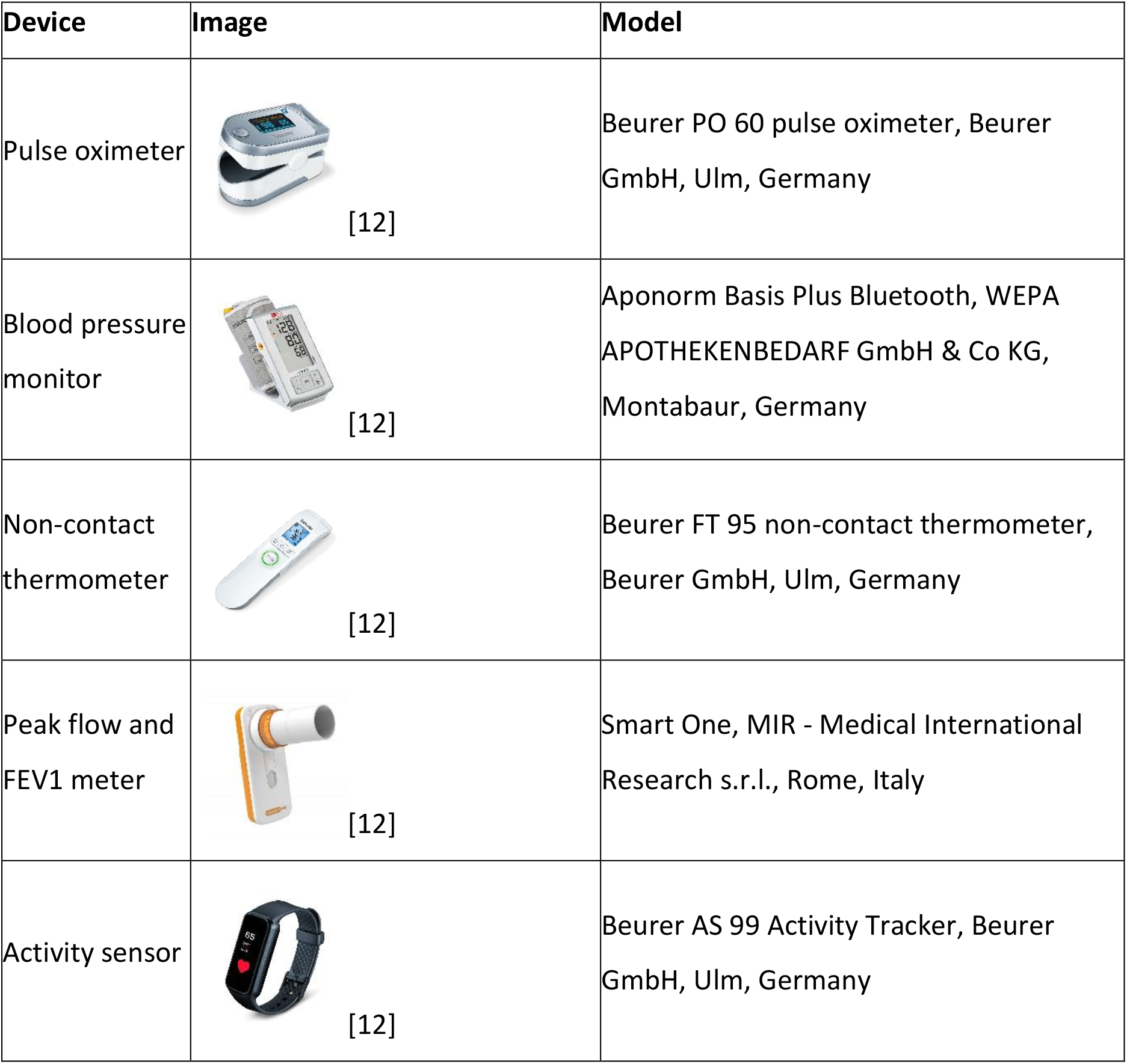
Measuring devices used in the study

| Device | Image | Model |
| --- | --- | --- |
| Pulse oximeter | [12] | Beurer PO 60 pulse oximeter, Beurer GmbH, Ulm, Germany |
| Blood pressure monitor | [12] | Aponorm Basis Plus Bluetooth, WEPA APOTHEKENBEDARF GmbH & Co KG, Montabaur, Germany |
| Non-contact thermometer | [12] | Beurer FT 95 non-contact thermometer, Beurer GmbH, Ulm, Germany |
| Peak flow and FEV1 meter | [12] | Smart One, MIR - Medical International Research s.r.l., Rome, Italy |
| Activity sensor | [12] | Beurer AS 99 Activity Tracker, Beurer GmbH, Ulm, Germany |

### Remote monitoring platform

By entering an authorization code, patients could link their account to their GP’s ‘SaniQ Praxis’ account, thus permitting them to view their data. Developments in their vital signs were displayed as a list or graph. GPs decided individually – i.e. according to a patient’s risk profile and the course of the disease - how often to check a patient’s data. Upper and lower limits for the vital signs were defined in accordance with standard operating procedures (SOPs) of participating pneumology departments and the COVID-19 treatment recommendations available at the time [15] (see supplementary material table S.1). GPs could also tailor these predefined limits to suit the needs of individual patients. If a vital sign was outside the limits, the patient’s GP received an automatic email from ‘SaniQ Praxis’ informing him or her accordingly. The GP could then check the available data on the platform and contact the patient if necessary. The optional alarm function could be deactivated by GPs for each individual patient.

### Data collection

#### a. Measurements

For 28 days, acute patients measured their heart rate, blood pressure, SpO_2_, and body temperature once a day. Post-acute patients determined the same vital signs for 12 weeks, supplemented by lung parameters (peak flow [PEF], forced expiratory pressure in 1 second [FEV1]) and activity parameters (daily step count).

#### b. Questionnaires

Patients filled in three in-app questionnaires consisting of questions relating to their medical history, daily symptoms and views on remote monitoring:

The medical history questionnaire included questions on current COVID-19 symptoms, relevant pre-existing medical conditions and current medication. Overall, the medical history questionnaire consisted of 35 questions requiring yes or no answers (see supplementary material table S.2) and was developed on the basis of available publications [16–19]. As the pandemic was evolving rapidly and studies in an ambulatory setting were urgently needed, the first patients were included while the questionnaire was still under development. The study team therefore took the medical history of the first 22 patients by phone, and filled in their medical history questionnaires retrospectively. In response to new research findings (e.g. skin rashes), further questions on symptoms were added later.

The second questionnaire consisted of seven questions on the most common COVID-19 symptoms. It was based on the SOPs of participating pneumology departments, DEGAM recommendations, and validated questionnaires on fatigue and shortness of breath [20–22]. Patients received the questionnaire daily for the duration of their participation in the study.

After completion of the study, patients were asked to fill in an evaluation questionnaire that asked about their experiences and satisfaction with remote monitoring (see supplementary material table S.3). The 11 questions were presented with a 4-point Likert scale and based on validated questionnaires on telemedicine satisfaction and telehealth usability [23–26]. Patients that stopped taking measurements before completing the study also received an evaluation questionnaire and, if necessary, a telephone or email reminder at the end of the study period.

### Data analysis

A certified server hosted in Germany was used as an online platform to store all data. Patient data were exported from the platform in a comma-separated values (csv) format. Personal data were pseudonymized before analysis. We used Microsoft Excel 2016 to descriptively analyze the data. Continuous variables such as age are provided with their mean and standard deviation, while absolute and relative frequencies are provided for discrete data.

The results also include information on the sample size n.

## 3. Results

### Sample Characteristics

Overall, 51 outpatients participated in the COVID-19@home study, of whom 32 were acute COVID-19 patients and 19 post-acute patients. Patients were between 19 and 77 years old, and their mean age was 48.7 (SD: 12.5). Thirty women (58.8%) and 21 men (41.2%) participated. Eight (15.7%) patients were obese (body mass index 30 or more). Information on further pre-existing conditions were provided by 48 patients (94.1%), of whom 10 (20.8%) had one risk factor for a potentially severe course of COVID-19, eight (16.7%) had two, and three (6.3%) had three risk factors. Eight patients had cardiovascular disease (16.7%), while six each had diabetes mellitus or lung disease (12.5%). The patients reported a mean of 4.2 symptoms upon study inclusion (SD: 2.8, minimum: 0, maximum: 11) (see table 2).

**Table 2.** COVID-19 symptoms (n=50)

| Questionnaire Item | Acute patients |  |  | Post-acute patients |  |  |
| --- | --- | --- | --- | --- | --- | --- |
|  | n (total)<sup>1</sup> | n<sup>2</sup> | % | n (total)<sup>1</sup> | n<sup>2</sup> | % |
| Number of symptoms upon study inclusion | 31 |  | 100 | 19 |  | 100 |
| • 0 |  | 0 | 0.0 |  | 7 | 36.8 |
| • 1-3 |  | 10 | 32.3 |  | 4 | 21.1 |
| • 4-6 |  | 13 | 41.9 |  | 6 | 31.6 |
| • 7-9 |  | 7 | 22.6 |  | 2 | 10.5 |
| • ≥10 |  | 1 | 3.2 |  | 0 | 0.0 |
| **Symptoms upon study inclusion** |  |  |  |  |  |  |
| Lethargy/exhaustion | 29 | 26 | 89.7 | 19 | 10 | 52.6 |
| Headache/dizziness | 10 | 8 | 80.0 | 19 | 7 | 36.8 |
| Cough | 31 | 23 | 74.2 | 19 | 4 | 21.1 |
| Pain in muscles/joints | 28 | 20 | 71.4 | 19 | 4 | 21.1 |
| Difficulty concentrating | 10 | 5 | 50.0 | 19 | 8 | 42.1 |
| Head cold | 29 | 12 | 41.4 | 19 | 3 | 15.8 |
| Sore throat | 30 | 12 | 40.0 | 19 | 3 | 15.8 |
| Gastro-intestinal complaints | 30 | 12 | 40.0 | 19 | 0 | 0.0 |
| Fever/chills | 31 | 11 | 35.5 | 19 | 1 | 5.3 |
| Smell/taste impairment | 30 | 11 | 36.7 | 19 | 6 | 31.6 |
| Breathing difficulties | 30 | 9 | 30.0 | 19 | 4 | 21.1 |
| Chest pain | 30 | 6 | 20.0 | 19 | 0 | 0.0 |
| Skin rash | 26 | 3 | 11.5 | 19 | 2 | 10.5 |

<sup>1</sup>n (total): number of patients being asked about symptoms. Differences in n (total) reflect regular updates to the medical history questionnaire.
<sup>2</sup>n: number of patients with symptoms.

### Participation

Acute patients started taking measurements 1 to 26 days after receiving their positive PCR test result, or an average of 5.7 days (SD: 5.7). Patients with post-acute symptoms began 22 to 447 days after receiving their positive PCR test result (mean: 152.4, SD: 112.6). Adherence to daily measuring was high, with 93.8% of acute patients and 89.5% of post-acute patients measuring vital signs as intended. The overall dropout rate was 7.8% (four patients).

### Evaluation

Overall, 48 patients received the evaluation questionnaire and 42 (87.5%) completed it. Of the respondents, 31 (73.8%) had ‘rarely’ or ‘never’ recorded their health data before. Nevertheless, 39 patients, or 92.9% ‘completely agreed’ or ‘partially agreed’ that they had managed to cope with using the app and measuring devices. All 42 respondents (100%) ‘completely agreed’ or ‘partially agreed’ that they ‘could comfortably integrate measurement-taking’ into their daily routines. Overall, 34 patients (81.0%) managed to deal with the app and devices on their own and ‘rarely’ or ‘never’ required aid in using them. Use of the app and measuring devices rarely seemed to have unsettled users: 38 (90.5%) of our cohort ‘partially disagreed’ or ‘disagreed’ that they had ‘experienced uncertainty when using the app and measuring devices’.

Twenty-five patients (59.5%) agreed that treatment of their COVID-19 illness had benefited from remote monitoring. Patients’ overall opinion of remote monitoring was very positive, with all 42 respondents describing it as ‘very good’ or ‘rather good’ (100%). Moreover, 39 of them (92.9%) would recommend the app and measuring devices to their family and friends (see table 3 for more details).

**Table 3.**
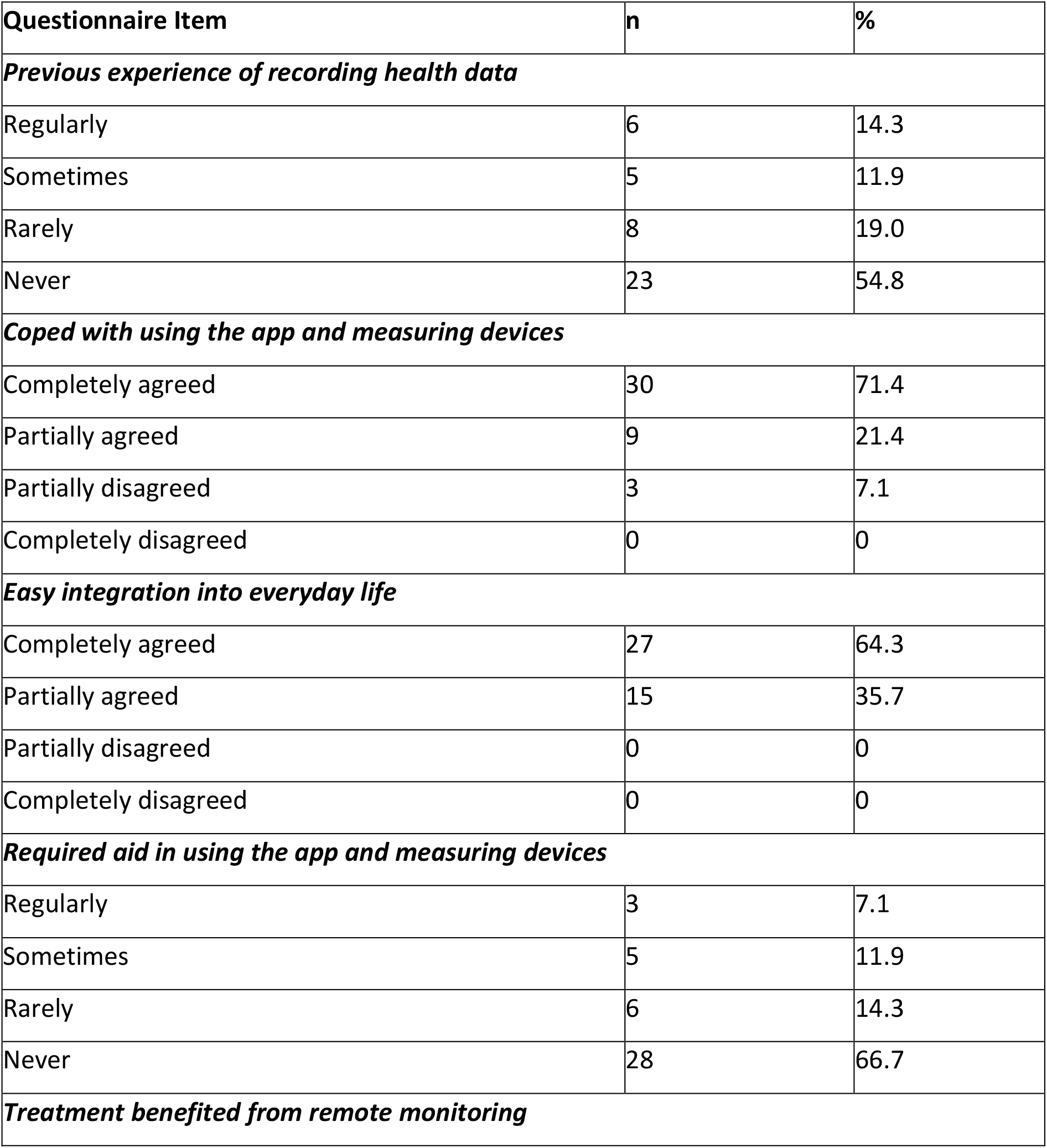
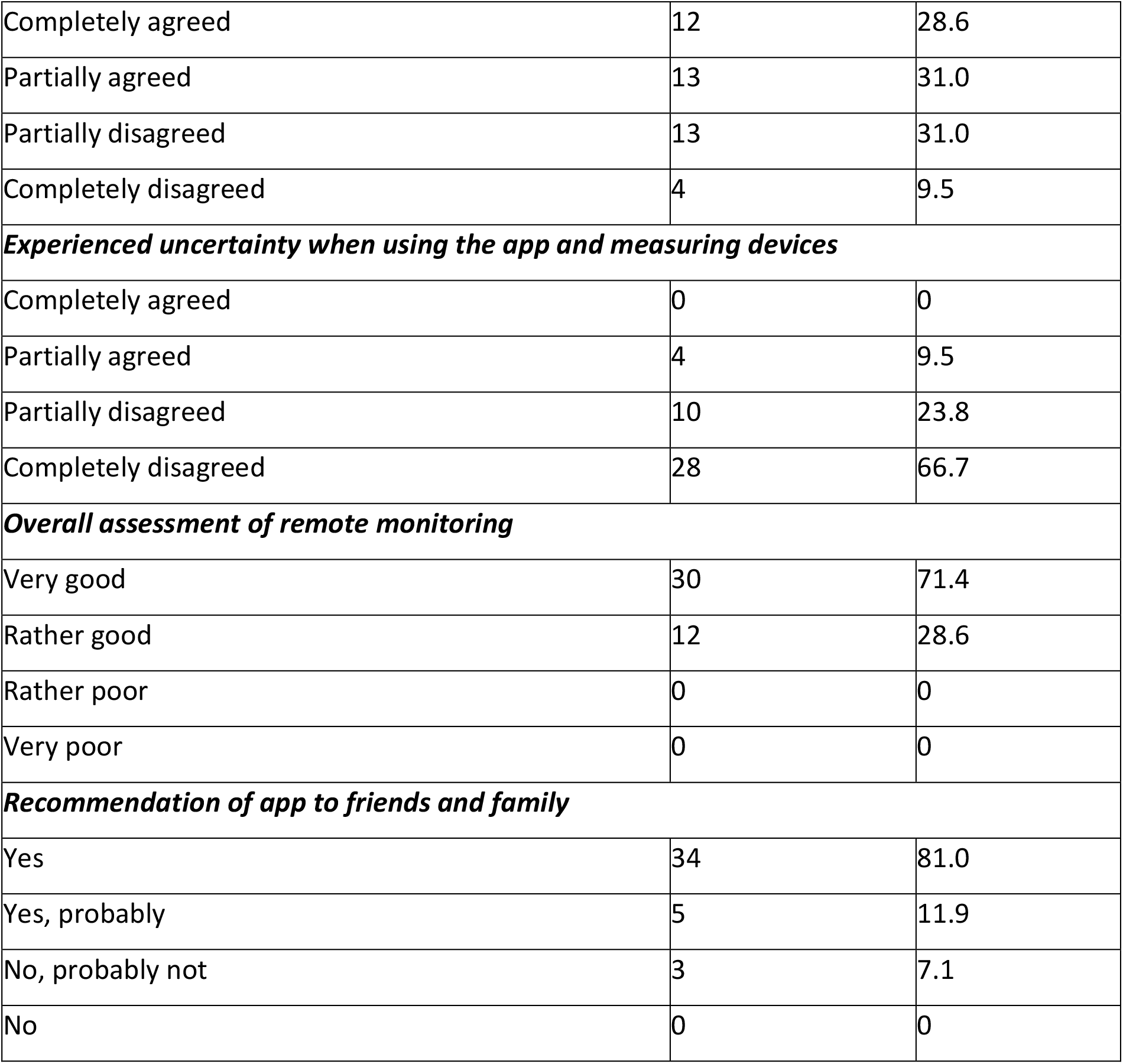
Evaluation of ‘SaniQ’ app and measuring devices (n = 42)

### Changes to the implementation process

In order to include acute patients as quickly as possible the study team put on full protection gear to avoid infection and handed over the consent forms and measuring devices in person. Afterwards, consent forms were returned via email. The measuring devices were sent to patients via express mail, as this proved to be reliable and fast enough (delivery within 48 hours).

We also distributed a detailed manual provided by the software partner, which resulted in a sharp drop in the number of calls due to technical problems.

As the pandemic was evolving rapidly and studies were urgently needed in an ambulatory setting, we started enrolling patients in January 2021, even though the development of our in-app questionnaires had not yet been finalized. During the pilot phase, we took patients’ medical histories and gathered information on their COVID-19 symptoms by telephone.

Further questions on symptoms were added when new research findings were published (e.g. gastro-intestinal complaints, skin rashes). The evaluation questionnaire was presented to 48/51 patients.

## 4. Discussion

### Main findings

We implemented a remote monitoring concept for COVID-19 outpatients. Patients used the included app to self-monitor their vital parameters and symptoms daily. GPs could then view the results on a software platform. Patient adherence was high with 93.8% of acute patients and 89.5% of post-acute patients measuring vital signs as intended. All patients that completed an evaluation questionnaire rated the app as ‘very good’ or ‘rather good’. Almost all patients would recommend the app and measuring devices to their families and friends. We optimized the implementation process during the pilot phase.

### Findings in relation to the literature

Our study is one of only a few to focus on patients’ use of remote monitoring to communicate with their GP practice [27]. In contrast to other studies and in order to gain broad experience, we included patients of all ages and with any pre-existing conditions. Several studies on remote monitoring have, however, investigated COVID-19 patients that required ongoing supervision and treatment in secondary settings, such as pre-admission wards, virtual wards, or that were receiving care in a ‘hospital at home’ (following an early release from hospital) [9, 28, 29]. Some studies included all patients suspected of having COVID-19, regardless of their PCR test results [30]. In our study, however, a positive PCR result was an inclusion criterion, which allowed us to focus on COVID-19 patients and their specific needs (e.g., self-isolation at home).

International studies on remote monitoring generally used pulse oximetry, while only a few used non-contact thermometers and blood pressure monitors [9, 28]. Telephone-, app- and paper-based protocols were used for data transfer [9, 28, 31, 32]. We decided to use an app to ensure instant documentation and data transfer, a low risk of transmission errors, and greater opportunities for data analysis. We managed to enrol acute COVID-19 patients quickly, and included them in our study a median of 3.5 days after a positive PCR test result. While other studies also emphasized the importance of speed, they provided no exact enrolment data [9].

Several authors have concluded that remote monitoring cannot be successful unless patients’ needs are taken into account [7, 28]. A lack of patient training, technical barriers, and insufficient usability appear to explain most patient dropouts [11, 28, 33]. In our study, some patients experienced initial difficulties installing the app and connecting the measuring devices. We therefore added a detailed manual to emails and packages containing the devices. This resulted in a sharp drop in the number of calls due to technical problems, and the overall dropout rate was low (7.8%). Patients’ evaluations indicate that our remote monitoring concept is suitable for patients that have no experience of recording health data. However, patients without German language skills could not participate in our remote monitoring program, unlike some remote home monitoring models in the UK that developed culturally appropriate patient information in different languages [28].

Our patients were advised to monitor their vital signs ‘at least once a day’. However, the frequency with which they measured them varied. This complicated data management, analysis and interpretation. We would therefore recommend specifying frequency more precisely in future studies. It might also be appropriate to adjust the frequency of measurements over the course of the disease and to intensify it in high-risk acute COVID-19 patients, as was the case in a study on remote monitoring conducted in Tuscany [34]. To raise compliance, post-acute patients might also be asked to take their measurements less often, e. g. three times a week.

### Strengths and limitations

Despite the relatively small sample size, our study provides real-world experience from patients who used the app intensively over a long period of time. They were thus able to accumulate considerable experience of remote monitoring, which increased the informative value of our evaluation results.

However, our study also has several limitations. As our questionnaires were still under development when the pilot phase began, they were partially conducted on the telephone and partially via the app itself. Furthermore, the medical history questionnaire evolved throughout the study to take account of new scientific findings on COVID-19 symptoms, limiting the generalizability of our results. In addition, we did not use a validated questionnaire, and several adjustments were made to the implementation process during the course of the study. At the beginning of the study, for example, we wore full protective gear to avoid infection and provided the consent forms and measuring devices in person. Afterwards, consent forms were distributed and collected via email, and measuring devices were sent to patients using express mail. As a result, the processes assessed by the patients were not identical, which may have limited the validity of our results.

However, as these modifications mainly concerned organizational processes, and the self-monitoring process remained unchanged, we would expect their influence on the evaluation to have been small. As quarantine limited technical support options, our approach to remote monitoring was only suitable for patients that owned a smartphone and were familiar with installing and using apps. It was therefore impossible to include persons that were unable to use the technical infrastructure. This also applied to people with language barriers or disabilities that would have prevented them from using the app and devices. In future research, attempts should be made to make remote monitoring possible for these people, too. Future research should also address cost-effectiveness and changes in such clinical outcomes as hospitalization and mortality, which we did not consider.

## 5. Conclusion

In this study, we implemented remote monitoring among COVID-19 outpatients and iteratively optimized our approach during a pilot phase. During the implementation process, we learned that the success of remote monitoring depends on certain conditions being fulfilled. First, non-contact delivery of measuring devices should be arranged for acute COVID-19 patients as soon as possible after the disease is diagnosed. Second, patients need precise and comprehensive instructions on how to correctly install and use the app and measuring devices. Patients’ evaluations indicate that most patients then accepted and were able to use remote monitoring as we intended. Third, as some patients initially experienced technical difficulties, background support was required to ensure both patient adherence and that the system worked as planned. However, it is difficult to imagine how general practices could cope with these additional tasks. Sufficient patient and practice support should therefore be provided if the model is to be implemented on a larger or even nationwide scale.

## Data Availability

All data produced in the present study are available upon reasonable request to the authors.

## Acknowledgments

The authors would like to express their thanks to all participating patients and GPs. We are also grateful to Phillip Elliott for the language review of the paper.

## Funding

This research was funded by the German Federal Ministry of Education and Research, BMBF, grant number 01KX2021.

## Competing interest statement

The institutions of S.H., S.M.K., P.J.C., N.D., L.N., T.E., R.G., R.K., G.R., M.D., C.F.V. and B.S.M received funding for this study from the German Federal Ministry of Education and Research, BMBF, grant number 01KX2021. The funder had no role in the design of the study; in the collection, analyses, or interpretation of data; in the writing of the manuscript, or in the decision to publish the results. P.J.C. is an employee at OncologyInformationService e.K. N.D. has received the CSL Behring award ‘Translational Research on Extracellular Vesicles in COPD’. R.K. and R.G. have been involved in projects and publications using the Kaia COPD App. SK is founder and managing partner of MED.digital GmbH. He holds shares of BioNTech, Pfizer, and CRISPR Therapeutics. He is member of the commission ‘New Professions’ of Stiftung Münch and of several committees of the German Medical Association (‘Digitalization of health care’ and ‘Medical Education and University Medicine’). B.S.M. has received personal honoraria for scientific consultancy to health insurance fund ‘Die Techniker’. The remaining authors declare no conflict of interest.

## Availability of data and materials

The datasets generated and analyzed during the study are available from the corresponding author on reasonable request.

## Notes

### Clinical Trial

German Clinical Trial Register ID (DRKS00024604, https://www.drks.de/drks_web/setLocale_EN.do, 26.04.2021)

### Funding Statement

This study was funded by the German Federal Ministry of Education and Research, BMBF, grant number 01KX2021.

### Author Declarations

Ethics committee of Goethe Universiy Frankfurt gave ethical approval for this work (No. 20-1023, 18.01.2021)

