## Supplementary Tables for "Implementing remote monitoring for COVID-19 patients in primary care"

### Supplementary Material

**Table S.1.** Upper and lower limits for the vital signs

| **Measurement** | **Limit[1]** |
| --- | --- |
| ***Patients with acute COVID-19*** | |
| Body temperature | ≤ 35.0°C or ≥ 39.0°C |
| Oxygen saturation (SpO2) | ≤ 93% |
| Systolic blood pressure | ≤ 90mmHg |
| Heart rate | ≤ 40/min or ≥ 110/min |
| ***Patients with post-acute COVID-19*** | |
| Body temperature | ≥ 39.0°C |
| Oxygen saturation (SpO2) | ≤ 93% |
| Systolic blood pressure | ≤ 90mmHg |
| Peak Expiratory Flow (PEF) in litres/min | Limits depend on gender, age and body size. Calculated using the 'SaniQ' app. |
| Forced Expiratory Pressure in 1 Second (FEV1) in litres | Limits depend on gender, age and body size. Calculated using the 'SaniQ' app. |

[1] based on standard operating procedures (SOPs) of participating pneumology departments and COVID-19 treatment recommendations available at that time [15].

**Table S.2.** COVID-19 symptoms at study inclusion: questionnaire in English

| **Questionnaire Item** | **Response Options** |
|---|---|
| Do you have fever or chills? | yes/no |
| Do you have a cough, with or without phlegm? | yes/no |
| Do you have difficulty breathing (shortness of breath, breathlessness, wheezing)? | yes/no |
| Do you feel lethargic or exhausted? | yes/no |
| Do you have difficulties concentrating? | yes/no |
| Is your sense of smell or taste impaired? | yes/no |
| Do you have a sore throat? | yes/no |
| Do you have a cold? | yes/no |
| Do you have a headache or feel dizzy? | yes/no |
| Do you have any pain in your muscles or joints? | yes/no |
| Have you any pain in your chest? | yes/no |
| Do you have any gastro-intestinal problems (diarrhea, vomiting, nausea)? | yes/no |
| Do you have a skin rash? | yes/no |

**Table S.3.** Evaluation of app and measuring devices: questionnaire in English

| Questionnaire Item | Response Options |
|---|---|
| I have taken notes of data relating to my health (e.g. blood pressure, or blood sugar, headache diary) in the past. | Regularly<br>Sometimes<br>Rarely<br>Never |
| I have no difficulty using the app and the measuring devices. | Completely agree<br>Somewhat agree<br>Somewhat disagree<br>Completely disagree |
| I could comfortably integrate measurement-taking into my daily routine. | Completely agree<br>Somewhat agree<br>Somewhat disagree<br>Completely disagree |
| Somebody helped me use the app and measuring devices. | Regularly<br>Sometimes<br>Rarely<br>Never |
| I have the feeling that the treatment of my COVID-19 illness benefited from remote monitoring. | Completely agree<br>Somewhat agree<br>Somewhat disagree<br>Completely disagree |
| I found the use of the app and measuring devices unsettling. | Completely agree<br>Somewhat agree<br>Somewhat disagree<br>Completely disagree |
| What is your overall opinion of remote monitoring? | Very good<br>Rather good<br>Rather poor<br>Very poor |

| | |
|---|---|
| Would you recommend use of the app and the measuring devices to your family and friends? | Yes<br>Yes, probably<br>No, probably not<br>No |
